# Estimating the importation risk of mpox virus in 2022 to Hong Kong, China

**DOI:** 10.1101/2023.03.17.23287412

**Authors:** Mingda Xu, Songwei Shan, Zengyang Shao, Yuan Bai, Zhanwei Du, Zhen Wang, Chao Gao

## Abstract

International air travel has been recognized as a crucial factor in the cross-regional transmission of monkeypox (now known as mpox) since this disease rapidly spread across the globe in May 2022. On September 6, 2022, Hong Kong SAR (HK) reported its first imported mpox case with travel history of the United States (US), Canada, and the Philippines. In this study, we estimated the importation risk to HK from 25 international departures from May 1 to September 6, 2022, based on the prevalence of pre-symptomatic mpox cases in the study regions, and time-varying flight mobility evaluated by aggregating multiple open-access air travel datasets (e.g., OpenSky, Aviation Edge). The result indicated that the US had the most significant importation risk of 0.63 (95% CI: 0.32, 0.95) during the study period, followed by the United Kingdom (UK) and Canada with a risk of 0.29 (95% CI: 0.10, 0.63) and 0.17 (95% CI: 0.08, 0.32), respectively. Our study demonstrated that the importation risk of mpox from the US and Canada was substantially higher than other regions, which was aligned with the travel history of the first reported case in HK. Our study provided a simplified computational method for estimating the importation risk of mpox virus based on air travel mobility and disease prevalence. Estimating the international importation risk of mpox is essential for appropriately designing and timely adjusting emergency public health strategies and inbound measures.

## Introduction

Mpox (monkeypox) is a zoonotic disease rarely transmitted outside Africa before 2022 (*Nature* 2022; S. Wang et al. 2022; Z. Du et al. 2022). On May 7, 2022, a confirmed case of mpox who traveled from Nigeria to the United Kingdom (UK) (“Monkeypox - United Kingdom of Great Britain and Northern Ireland” n.d.) was reported by the World Health Organization. After that, the rapid outbreak marked the first time that mpox spread widely in non-endemic countries, covering Europe, North America, and Oceania successively (“Multi-Country Monkeypox Outbreak in Non-Endemic Countries” n.d.). On July 23, 2022, World Health Organization declared the ongoing mpox outbreak as a Public Health Emergency of International Concern (PHEIC) (“WHO Director-General Declares the Ongoing Monkeypox Outbreak a Public Health Emergency of International Concern” n.d.). As of December 28, 2022, 83,751 cases had been reported in 110 countries or regions, with 9 countries reporting a high cumulative number of cases (≥3000), namely the US (n=29,554), Brazil (n=10,493), Spain (n=7,496), France (n=4,114), Colombia (n=4,021), UK (n=3,730), Germany (n=3,676), Peru (n=3,643), and Mexico (n=3,509) (Mathieu et al. 2022).

International air travel plays an essential role in the transmission of mpox (Talbot 2022). Most confirmed cases were with travel history from European and North American countries rather than West or Central Africa (“Monkeypox Outbreak” n.d.; Angelo et al. 2019). The first imported mpox case in Hong Kong SAR (HK), whose recent stops included the US, Canada, and the Philippines, was documented on September 6, 2022 (*Reuters* 2022a). The Government of Hong Kong has procured a third-generation vaccine, “JYNNEOS,” which has been licensed by the U.S. Food and Drug Administration (FDA) to protect against mpox (“Centre for Health Protection, Department of Health - Monkeypox” n.d.). Although rapid responses were quickly initiated in non-endemic areas, including vaccination and surveillance of human-to-human transmission, travelers from high-risk regions may still pose a potential importation risk to the destinations due to the long incubation period (Thornhill et al. 2022).

Various mathematical models and research methods have been developed to estimate the risk of cross-regional transmission. Previous research has provided vital insight into infectious disease transmission by combining travel data with local disease outbreak data (Shi et al. 2020; Kuo and Chiu 2021; Russell et al. 2021; Zhang et al. 2022). In the fight against COVID-19 outbreaks, the importation risk of COVID-19 (including the wildtype, 501Y, and Omicron sub-variants) is quantified based on the accessibility to the airline network and local disease situations to guide inbound control measures (Z. Du et al. 2021; Bai, Du, et al. 2022; Nakamura and Managi 2020; Z. Du et al. 2020; Bai, Xu, et al. 2022; Xu et al. 2022). The correlations between air travel and the international transmission of mpox have been clearly identified (Bhattacharya, Dhama, and Chakraborty 2022; M. Du et al. 2022; Kinoshita et al. 2022). These studies highlighted the importance of appropriately estimating the importation risk among air passengers to effectively mitigate the cross-regional transmission of infectious disease (Menkir et al. 2021; Lai et al. 2022; Han et al. 2021).

As a major international transportation hub and a special economic zone of China, HK has been affected by the global spread of infectious diseases over the past decades (Z. Du, Tian, and Jin 2022; Z. Du et al. 2023). HK experienced the SARS outbreak in 2003 (Pine and McKercher 2004) as well as the Influenza A (H1N1) pandemic in 2009 (Liao et al. 2010), and further fueled and accelerated the global transmission of these diseases (National Immunization Advisory Committee (NIAC) Technical Working Group (TWG), Influenza Vaccination TWG 2021). The volume of air traffic from overseas to Hong Kong has been gradually increasing following the reopening of HK on August 12, 2022 (“Government Announces Lifting of Compulsory Quarantine Requirement on Arrival at Hong Kong” n.d.), understanding and nowcasting the international importation risk of the mpox virus is critical for designing appropriate emergency public health strategies and inbound measures to reduce transmission risk. Our study aims to quantify the importation risk of mpox by combining aviation transportation data and prevalence in origin regions estimated during the early stage of the emergency public health outbreak, using HK as a case study. We estimated the daily volumes of air travel passengers from international regions to HK via open-access aviation datasets (e.g., OpenSky (Meides n.d.), Aviation Edge (“Aviation API List” 2017)) and the prevalence of mpox by country from Our World In Data (Mathieu et al. 2022). We further estimated the probability of importing at least one mpox case between May 1 and September 6, 2022, before the date of the first mpox imported case. Our research included 25 regions covering four continents with the most reported cases and direct air connection to HK.

## Materials and Methods

### Data Source

To estimate the daily number of passengers traveling to HK, we aggregated the daily direct flights arriving (and landing) in HK by combining the results from the OpenSky (Meides n.d.) and Aviation Edge (“Aviation API List” 2017). The information for each airport and estimated passenger capacity per aircraft were obtained from OurAirports (“Open Data @ OurAirports” n.d.) and the Admtl (Aéroports de Montréal 2021), respectively.

We retrieved monthly data for about 696 thousand air-travel passengers arriving or transiting in HK during the study period ***T*** (from May 1 to September 6, 2022) (“Tourism Statistics” n.d.). In total, we extracted 20,181 direct and transit flights from 46 distinct regions, with 14,858 from OpenSky and 5,323 additional from Aviation Edge. Due to the lack of transit flight information, we assumed Hong Kong was the destination for every flight landing at Hong Kong International Airport. We obtained the 7-day smoothed daily reported cases of mpox of different countries and regions from Our World In Data (Mathieu et al. 2022) from May to September 2022.

### Estimating daily air-travel passengers from international regions to HK

We first estimate the time-varying mobility via air travel following the methods used in ref. (Bai et al. 2022) and then calculate the prevalence of pre-symptomatic cases in a given area to estimate the likelihood of introductions. The notation and values of the parameters are provided in **Table 1**.

**Table 1:**
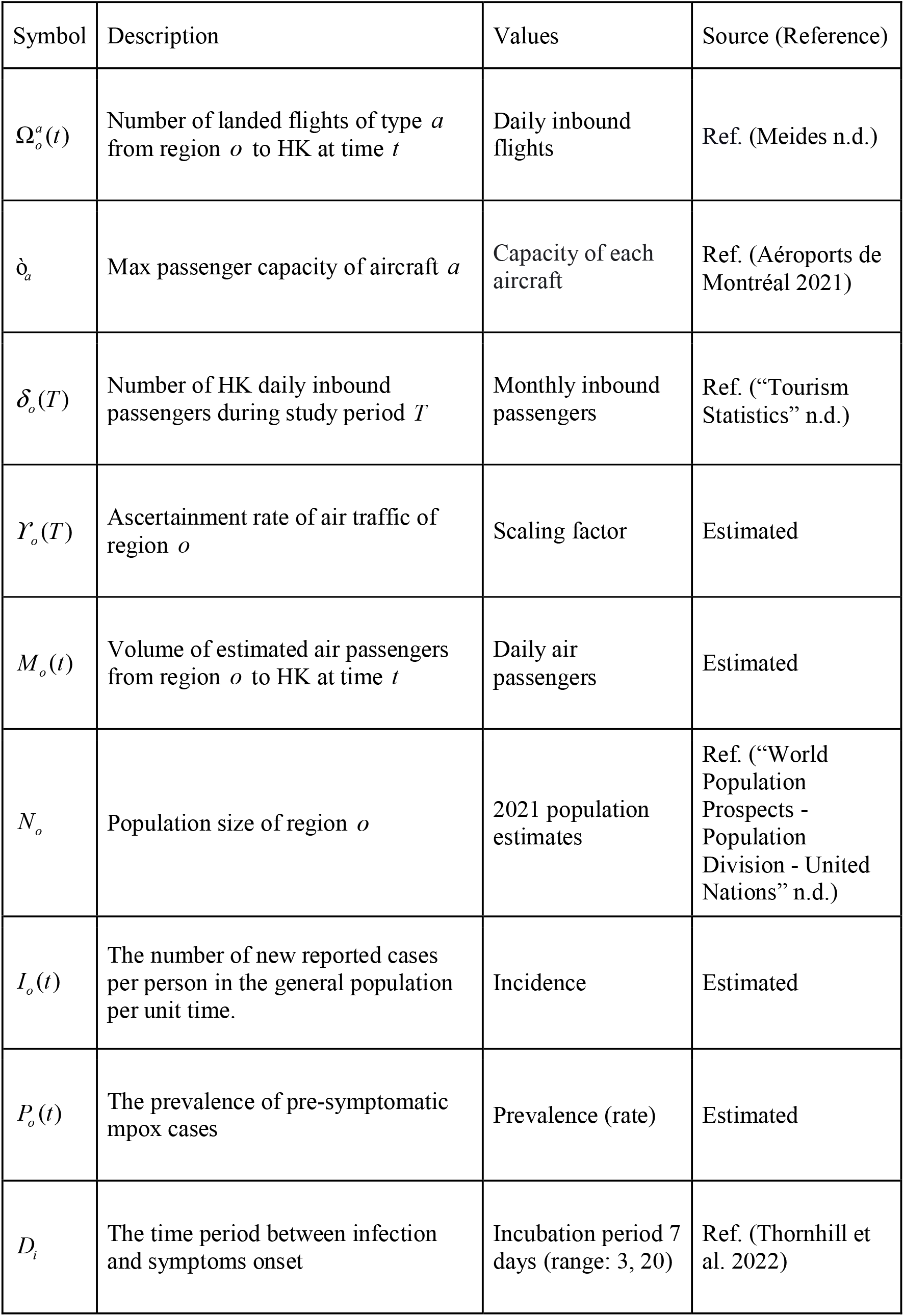
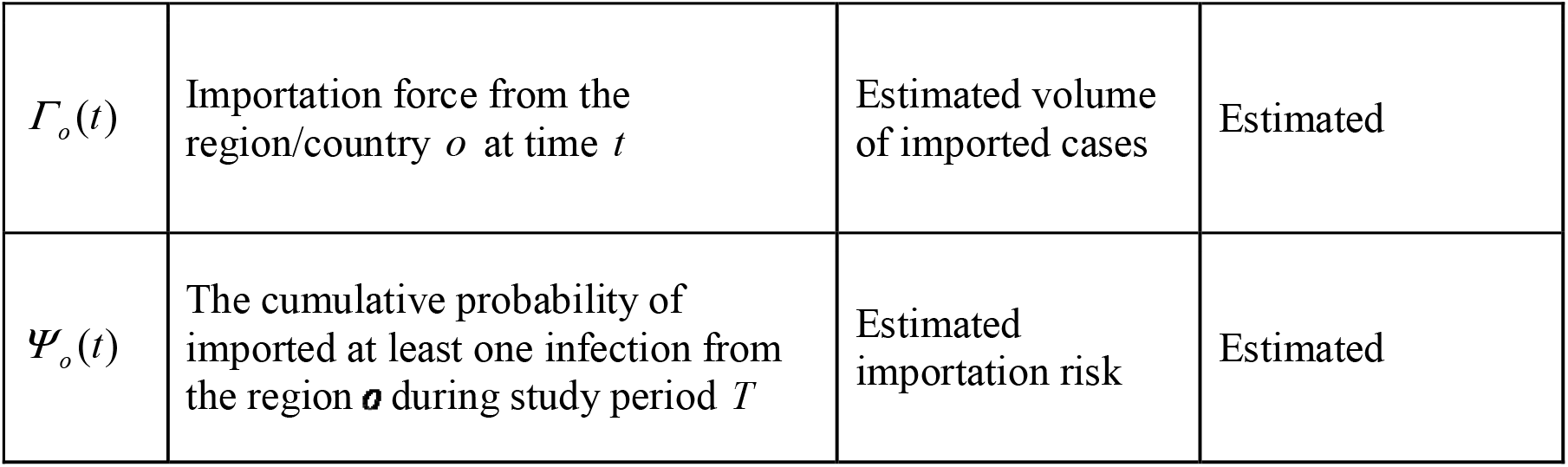
Model Parameters and Data Sources.

Given the lack of information regarding the actual composition of each flight, it was assumed that each flight was at full capacity. Used 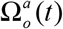 is the daily volume of aircraft ***a*** from the international region *o* to HK and ò_*a*_ is the maximum passenger capacity for aircraft *a* of all types of aircraft *A* (Aéroports de Montréal 2021). Thus, the approximated inbound passengers *m*_*o*_(*t*) from *o* to HK on day *t* is given by: 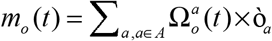 Then, we validated the air traffic by defining the ascertainment rate of passenger volume *ϒ*_*o*_(*T*) as the ratio of actual and estimated passengers to adjust the probably overestimated air-travel passengers, given by:

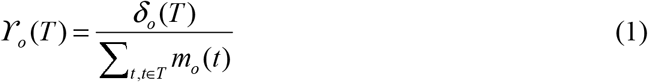

where *δ*_*o*_(*T*) stands for the actual passengers from the *o* during *T*. For those regions without the monthly visitor statistics to HK (“Tourism Statistics” n.d.), we matched *m*_*o*_(*t*) and the ascertainment rate *ϒ*_*o*_(*T*) to estimate the daily inbound volume: *M*_*o*_(*T*) = *ϒ*_*o*_(*T*) × *m*_*o*_(*t*).

### Estimating pre-symptomatic prevalence of mpox

We utilized 7-day smoothed mpox cases reported by region (Mathieu et al. 2022) to estimate the pre-symptomatic prevalence of mpox. Following the sub-exponential model (Chowell et al. 2016; Z. Du et al. 2022), we first estimated the confirmed case on day ^*t*^ during the early outbreak of epidemics by:

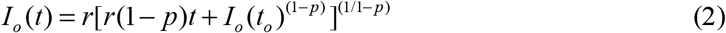

where *t*_0_ indicates the date when the case was first detected in region *o* and *I*_*o*_(*t*_*o*_) the corresponding initial number, *r* and *p* are the growth rate and the deceleration of growth, respectively. We assume *r* and *p* follow the normal distributions and estimate their mean and standard variance based on the daily confirmed cases in the study region, using the Levenberg–Marquardt algorithm implemented in prior studies (Viboud, Simonsen, and Chowell 2016) to minimize the residual sum of squares (RSS) between the model output and the observed cases.

To calculate the total number of infected cases who had not yet developed symptoms *Φ*_*o*_(*t*), we back-shifted the time series of reported cases by incubation period *D*_*i*_:

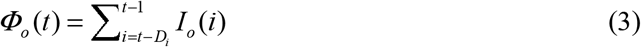

Then, we divided the total cases by the population size *N*_*o*_ of the origin **o** to calculate the prevalence of pre-symptomatic cases, which is given by *P*_*o*_(*t*) = *Φ*_*o*_ (*t*) / *N*_*o*_. For each simulation to yield the incidence in the region *o* during the study period, we independently simulate 1000 times to sample a pair of *r* and *p* from the normal distributions to introduce uncertainty.

### Estimating importation risk of mpox

We assumed all the mpox cases (that had not developed symptoms among passengers who intended to travel to HK) would be able to board. To estimate the number of imported cases from each region, we combined air-passenger with the daily prevalence of mpox to yield the importation force of cases that have not developed symptoms. We assumed that the proportion of pre-symptomatic passengers departing from a region was the same as the overall prevalence of pre-symptomatic cases in that region. Thus, the imported cases from region *o* to HK on day ***t***, is given by:

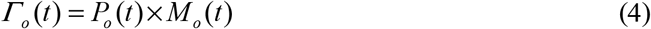

Informed by the first case among inbound passengers reported in HK who might be infected in the US on September 6, 2022 (*Reuters* 2022a), we calibrated the cumulative number of imported cases from the US as 1 at the end of the study period. Based on the observed event by September 6, 2022, we normalized each origin region’s cumulative imported cases, by 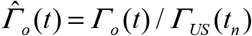, where *Γ*_*US*_(*t*_*n*_) = 1 denotes the observed imported cases from the US on ***t***_***n***_ (September 6, 2022).

Assuming that the imported cases of mpox from the study region ***o*** to HK are essentially a non-homogeneous poisson process (Wang and Wu 2018; Silverstein et al. 2020), we estimated the cumulative probability of at least one infected case (Z. Du et al. 2021; Bai et al. 2022) being introduced from ***o*** to HK by *t* as:

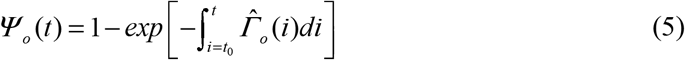

We estimated the time-varying flight mobility from a given country to HK with multiple open-access air travel datasets and the prevalence of pre-symptomatic cases in each study region. We synthesized these results to calculate the likelihood of at least one imported case occurring before September 6, 2022. Two indicators for the international importation risk of mpox to HK, namely the cumulative volume of imported cases and the cumulative probability of importing at least one case, were estimated by combining the aviation transportation data and the prevalence in origin regions The estimated importation risk for the 25 regions between May 1 and September 6, 2022 is shown in **Table 2**.

**Table 2:** Importation risk estimated during the study period. We estimated the cumulative risk of importation (95% CI) from 25 regions as of September 6, 2022. The region’s name in bold denotes the first case detected in HK who arrived from the Philippines after traveling in the US and Canada.

| Location | Importation Risk (95% CI) |
| --- | --- |
| **United States** | 0.63212 (0.32408, 0.94727) |
| United Kingdom | 0.29169 (0.09799, 0.62705) |
| **Canada** | 0.17269 (0.07545, 0.31758) |
| France | 0.09210 (0.03152, 0.24752) |
| Germany | 0.08175 (0.02088, 0.24205) |
| Netherlands | 0.05098 (0.01608, 0.12905) |
| Luxembourg | 0.01815 (0.00942, 0.03613) |
| Australia | 0.01730 (0.01035, 0.03053) |
| Singapore | 0.01702 (0.00772, 0.03668) |
| Belgium | 0.01045 (0.00481, 0.02068) |
| Switzerland | 0.00504 (0.00282, 0.00856) |
| Israel | 0.00446 (0.00254, 0.00761) |
| Italy | 0.00323 (0.00141, 0.00732) |
| Qatar | 0.00240 (0.00125, 0.00438) |
| New Zealand | 0.00114 (0.00061, 0.00214) |
| United Arab Emirates | 0.00096 (0.00012, 0.00362) |
| Thailand | 0.00063 (0.00041, 0.00097) |
| Finland | 0.00038 (0.00013, 0.00115) |
| **Philippines** | 0.00011 (0.00004, 0.00019) |
| Japan | 0.00010 (0.00006, 0.00020) |
| South Korea | 0.00009 (0.00002, 0.00019) |
| Saudi Arabia | 0.00007 (0.00005, 0.00010) |
| India | 0.00003 (0.00002, 0.00006) |
| Turkey | 0.00001 (0.00001, 0.00002) |
| Indonesia | 0.00001 (0.00000, 0.00001) |

**Figure 1a** shows the estimated number of imported cases in HK by September 6, 2022. The x-axis designates the estimated cumulative imported cases. Among the 25 international regions included in this study, we estimated that the US imported the maximum cases at 1.0 (95% CI: 0.39, 2.89), followed by UK at 0.35 (95% CI: 0.10, 0.99), Canada at 0.19 (95% CI: 0.08, 0.38).

**Figure 1:**
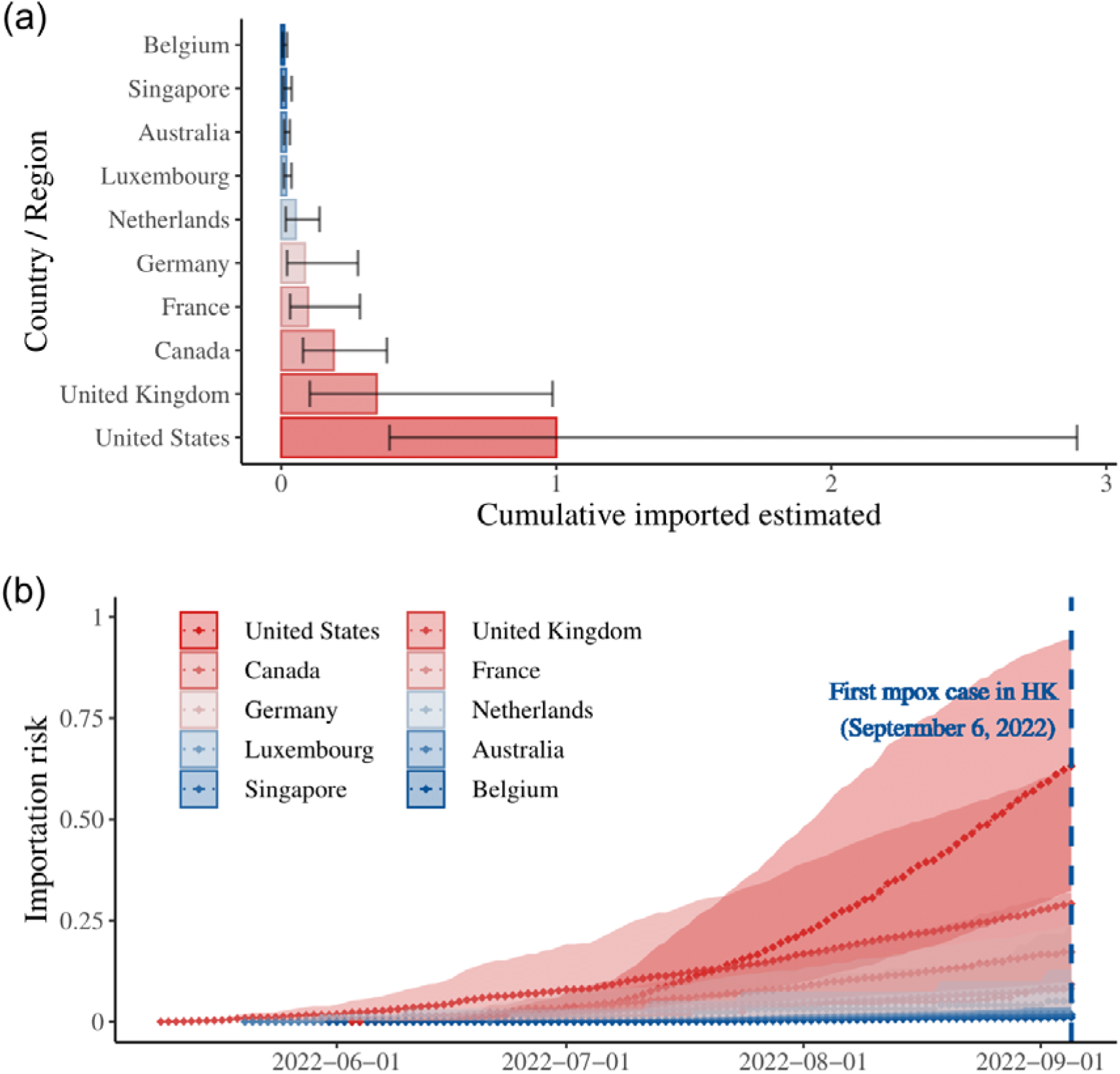
Estimated risk of mpox introductions from international regions to HK via 1000 stochastic simulations from May 1 to September 6, 2022. (a) As of September 6, 2022, the estimated number of imported cases arrived in Hong Kong based on OpenSky data and Aviation API database. The barplot denotes the estimated imported cases from the top 10 regions during the study period, with 95% CI; (b) The probability that at least one person infected with mpox to HK by the date indicated on the x-axis, the color of line corresponds to the estimated risk of each origin location as of September 6, 2022, with red and blue denoting high and low probability, respectively.

**Figure 1b** represents the curve of importation risk reflecting the cumulative probability of importing at least one mpox case among inbound travelers. Based on the observed sources of imported cases, only the US had a mean importation risk of 0.63 higher than 0.5 (95% CI: 0.32, 0.95) as of September 6, 2022. The UK and Canada are the following, with 0.29 (0.10, 0.63) and 0.17 (0.08, 0.32), respectively.

**Figure 2** displays the risk of importation of mpox for the study regions as of September 6, 2022, where the closer the color to red indicates a higher risk of importation in that region. Our study found that the risk from the US and Canada was relatively higher, which is aligned with the fact that the first reported case in Hong Kong had stayed in both countries before arriving and may have been infected there.

**Figure 2:**
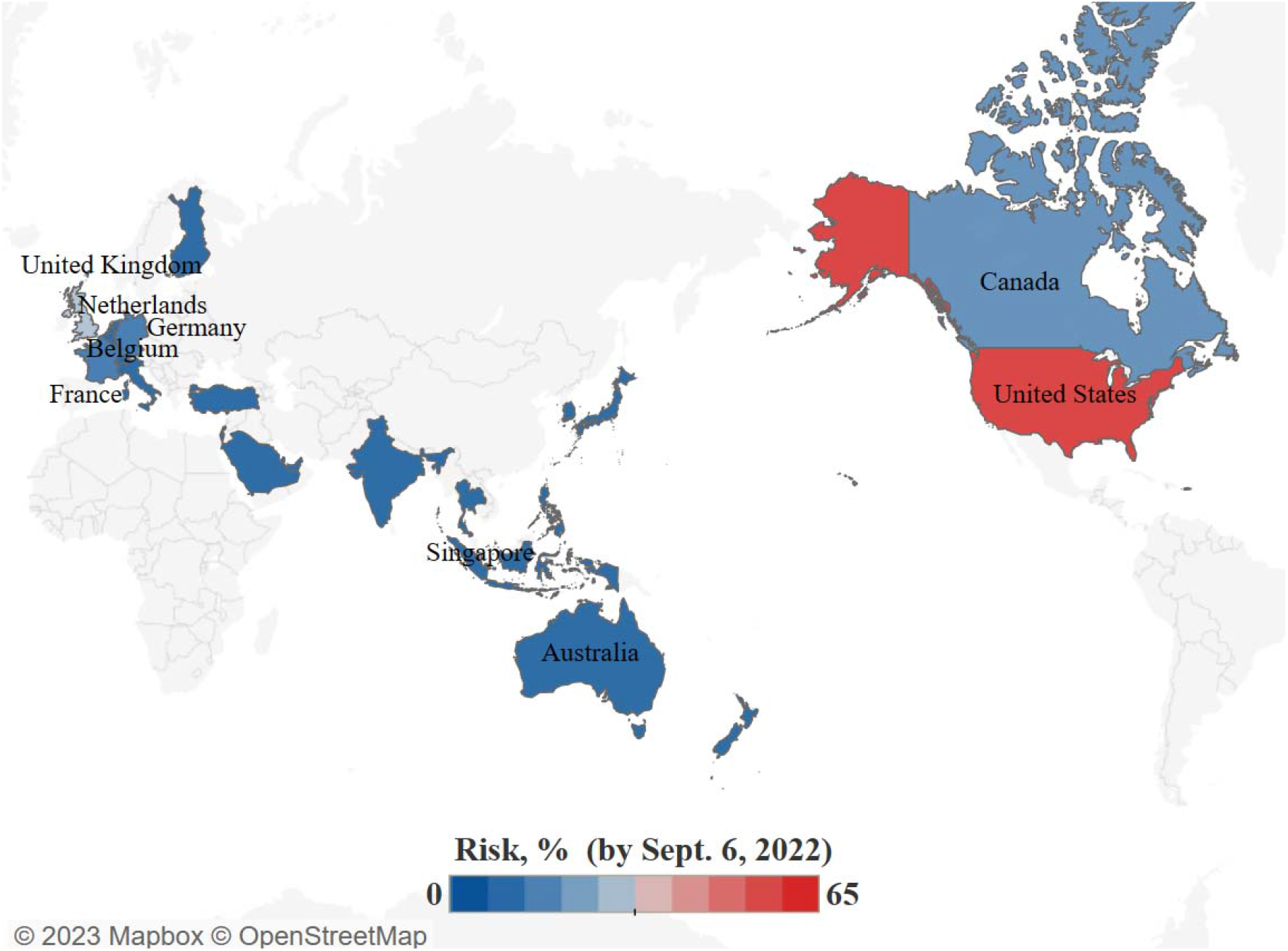
Risk map of cumulative probability of at least one mpox case imported from international regions by September 6, 2022 in 1000 stochastic simulations. Regions in red to blue indicate high and low risk. Regions in grey were not analyzed because mobility data were unavailable or no reported cases of mpox.

## Discussion

Human mobility patterns influence epidemiological risk (Gao and Liu 2013). The prevalence of a newly emerging infectious disease will most likely be determined by where it was first imported. In this study, we estimated the time-varying importation risk of mpox from 25 international regions to HK. Informed by the estimated arrival number of passengers and the local prevalence of mpox in each international origin, we developed a statistical method to estimate the cumulative risk of importation of mpox by combining actual imported events in HK. Our results were aligned with reality, and the estimates of imported cases reflected the connectivity of air travel and the prevalence of mpox in the corresponding origin countries and regions.

Our findings emphasize the importance of enhancing surveillance against mpox among travelers in regions with a high importation risk of mpox. The cumulative risk of importation of a region is highly correlated with the aggregated air travel flow to HK and the prevalence of mpox in that region. The ranking of importation risk indicated that the US had the highest risk during the entire study period. The number of mpox cases in the US surged from July to August (CDC 2022b), and the number of travelers from the US continuously increased during this period as the gradual relaxation of Hong Kong’s bounder control (“Tourism Statistics” n.d.). Except for the US, where the imported cases have been observed, several non-endemic countries, including the UK, Canada, France, Germany, and the Netherlands, are also at relatively high risk of importation. A previous study based on the Risk Matrix Method (M. Du et al. 2022) drew a similar conclusion about the importation risk of mpox into China: The US had the highest importation risk of mpox and should be closely monitored. Globally, MSM (men who have sex with men) make up the majority of cases in the current outbreak (CDC 2022a). The mpox virus is typically transmitted by close, personal contact with groups who developed symptoms among healthcare workers, laboratory personnel, and people with “high-risk sexual practices” such as sex workers (*Reuters* 2022b). Although the probability of importing a case of mpox via international aviation is extremely low at the end of August, implementing surveillance and contact tracing for passengers with high-risk activities from specific areas would effectively contain the cross-regional spread of mpox.

Several assumptions and limitations should be discussed in our estimations. First, due to no access to the complete commercial aviation dataset, we postulated that air travels between regions are all directed, while the indirect travels should also be included in the air traffic volumes. This may cause the incorrect estimation of the passenger volumes to HK. We expect that combining complete passenger data with flight transit information would improve the accuracy of estimates of potential importation risks from international departures, but would not substantially change the analysis results. Second, our method for estimating mpox prevalence relies on the number of reported cases in the early phase of an outbreak. Given the varieties in monkeypox surveillance capacity in different regions and the delay between infection and case reporting (“Monkeypox 2022 Global Epidemiology; Report 2022-09-23” n.d.), the resulting undetected cases may lead to underestimate local pre-symptomatic prevalence. Third, we assumed that only infected cases who had not developed symptoms could board following prior studies (Z. Du et al. 2021). While in reality, some individuals with asymptomatic or mild infection may also be able to go aboard (De Baetselier et al. 2022). Obtaining data on the proportion of mild and asymptomatic infection in the real world would improve our estimates of mpox importation risk among air travelers. Fourth, our study only examined the importation risks posed by air travelers, hence the results cannot be extrapolated to other transportation routines (Menkir et al. 2021; Gao et al. 2022).

## Conclusions

To conclude, our study provided a simplified computational method for estimating the mpox virus importation risk based on air travel mobility and disease prevalence. The approach can be adjusted to incorporate prevalence estimates and flight data from any origin and destination locations requiring minimum data. Our findings emphasize the necessity for a more comprehensive understanding of the probable sources of case importation for predictive simulation and risk perception. Reliable estimates of international importation risk from an endemic or non-endemic area will benefit in accurately evaluating the potential need to curtail the cross-regional transmission of the mpox virus as well as the implementation of disease monitoring and preparedness.

## Data Availability

The data underlying this article and these programs will be shared on reasonable request with the corresponding author (email to the corresponding author). The database of the air travel and mpox cases is publicly available at Crowdsourced air traffic data from The OpenSky Network 2020 | Zenodo, Aviation Edge - Database and API - Aviation database and API (aviation-edge.com), Mpox (monkeypox) - Our World in Data, respectively.

## Conflicts of Interest

The authors declare that there is no conflict of interest regarding the publication of this article.

## Author Contributions

MX, ZD, SS, ZS and YB: conceived the study, designed statistical and modeling methods, conducted analyses, and wrote the manuscript; CG, ZW: interpreted results and revised the manuscript.

## Funding Statement

This work was supported by the Key Program for International Science and Technology Cooperation Projects of China [grant no. 2022YFE0112300], National Natural Science Foundation of China [grant nos. 61976181, 62261136549, U22B2036], Key Technology Research and Development Program of Science and Technology - Scientific and Technological Innovation Team of Shaanxi Province [grant no. 2020TD-013].

## Supplementary Materials

There are no Supplementary Materials in this paper.

## Notes

### Competing Interest Statement

The authors have declared no competing interest.

## References

Aéroports de Montréal. 2021. “Reference Table – Number of Seats per Aircraft Type.” Admtl. 2021. https://www.admtl.com/sites/default/files/2021/2021_Appendix_6-Number-of-seats-per-aircraft-type.pdf.

Angelo, Kristina M., Brett W. Petersen, Davidson H. Hamer, Eli Schwartz, and Gary Brunette. 2019. “Monkeypox Transmission among International Travellers-Serious Monkey Business?” Journal of Travel Medicine 26 (5). https://doi.org/10.1093/jtm/taz002.

“Aviation API List.” 2017. Aviation Database and API. Aviation Edge. October 16, 2017. https://aviation-edge.com/aviation-api-list/.

Bai, Yuan, Zhanwei Du, Mingda Xu, M. Sc, Lin Wang, Peng Wu, Eric H. Y. Lau, Benjamin J. Cowling, and Lauren Ancel Meyers. 2022. “International Risk of SARS-CoV-2 Omicron Variant Importations Originating in South Africa.” Journal of Travel Medicine, June. https://doi.org/10.1093/jtm/taac073.

Bai, Yuan, Mingda Xu, Caifen Liu, Mingwang Shen, Lin Wang, Linwei Tian, Suoyi Tan, et al. 2022. “Travel-Related Importation and Exportation Risks of SARS-CoV-2 Omicron Variant in 367 Prefectures (Cities)— China, 2022.” China CDC Weekly 4 (40): 885–89.

Bhattacharya, Manojit, Kuldeep Dhama, and Chiranjib Chakraborty. 2022. “Recently Spreading Human Monkeypox Virus Infection and Its Transmission during COVID-19 Pandemic Period: A Travelers’ Prospective.” Travel Medicine and Infectious Disease 49 (June): 102398.

CDC. 2022a. “2022 Outbreak Cases and Data.” Centers for Disease Control and Prevention. November 21, 2022. https://www.cdc.gov/poxvirus/monkeypox/response/2022/index.html.

CDC. 2022b. “U.S. Mpox Case Trends Reported to CDC.” Centers for Disease Control and Prevention. December 14, 2022. https://www.cdc.gov/poxvirus/monkeypox/response/2022/mpx-trends.html.

“Centre for Health Protection, Department of Health - Monkeypox.” n.d. Accessed December 30, 2022. https://www.chp.gov.hk/en/healthtopics/content/24/101721.html.

Chowell, Gerardo, Cécile Viboud, Lone Simonsen, and Seyed M. Moghadas. 2016. “Characterizing the Reproduction Number of Epidemics with Early Subexponential Growth Dynamics.” Journal of the Royal Society, Interface / the Royal Society 13 (123): 20160659.

De Baetselier, Irith, Christophe Van Dijck, Chris Kenyon, Jasmine Coppens, Johan Michiels, Tessa de Block, Hilde Smet, et al. 2022. “Retrospective Detection of Asymptomatic Monkeypox Virus Infections among Male Sexual Health Clinic Attendees in Belgium.” Nature Medicine, August. https://doi.org/10.1038/s41591-022-02004-w.

Du, Min, Shi Mo Zhang, Wei Jing Shang, Wen Xin Yan, Qiao Liu, Chen Yuan Qin, Min Liu, and Jue Liu. 2022. “2022 Multiple-Country Monkeypox Outbreak and Its Importation Risk into China: An Assessment Based on the Risk Matrix Method.” Biomedical and Environmental Sciences: BES 35 (10): 878–87.

Du, Zhanwei, Zengyang Shao, Yuan Bai, Lin Wang, Jose L. Herrera-Diestra, Spencer J. Fox, Zeynep Ertem, Eric H. Y. Lau, and Benjamin J. Cowling. 2022. “Reproduction Number of Monkeypox in the Early Stage of the 2022 Multi-Country Outbreak.” Journal of Travel Medicine 29 (8). https://doi.org/10.1093/jtm/taac099.

Du, Zhanwei, Linwei Tian, and Dong-Yan Jin. 2022. “Understanding the Impact of Rapid Antigen Tests on SARS-CoV-2 Transmission in the Fifth Wave of COVID-19 in Hong Kong in Early 2022.” Emerging Microbes & Infections 11 (1): 1394–1401.

Du, Zhanwei, Lin Wang, Simon Cauchemez, Xiaoke Xu, Xianwen Wang, Benjamin J. Cowling, and Lauren Ancel Meyers. 2020. “Risk for Transportation of Coronavirus Disease from Wuhan to Other Cities in China.” Emerging Infectious Diseases 26 (5): 1049–52.

Du, Zhanwei, Lin Wang, Bingyi Yang, Sheikh Taslim Ali, Tim K. Tsang, Songwei Shan, Peng Wu, Eric H. Y. Lau, Benjamin J. Cowling, and Lauren Ancel Meyers. 2021. “Risk for International Importations of Variant SARS-CoV-2 Originating in the United Kingdom.” Emerging Infectious Diseases 27 (5): 1527.

Du, Zhanwei, Xiao Zhang, Lin Wang, Sidan Yao, Yuan Bai, Qi Tan, Xiaoke Xu, et al. 2023. “Characterizing Human Collective Behaviors During COVID-19—Hong Kong SAR, China, 2020.” China CDC Weekly 5 (4): 71–75.

Gao, Chao, Yi Fan, Shihong Jiang, Yue Deng, Jiming Liu, and Xianghua Li. 2022. “Dynamic Robustness Analysis of a Two-Layer Rail Transit Network Model.” IEEE Transactions on Intelligent Transportation Systems 23 (7): 6509–24.

Gao, Chao, and Jiming Liu. 2013. “Modeling and Restraining Mobile Virus Propagation.” IEEE Transactions on Mobile Computing 12 (3): 529–41.

“Government Announces Lifting of Compulsory Quarantine Requirement on Arrival at Hong Kong.” n.d. Accessed January 1, 2023. https://www.info.gov.hk/gia/general/202209/24/P2022092400048.htm.

Han, Xiaoyi, Yilan Xu, Linlin Fan, Yi Huang, Minhong Xu, and Song Gao. 2021. “Quantifying COVID-19 Importation Risk in a Dynamic Network of Domestic Cities and International Countries.” Proceedings of the National Academy of Sciences of the United States of America 118 (31). https://doi.org/10.1073/pnas.2100201118.

Kinoshita, Ryo, Miho Sassa, Shogo Otake, Fumi Yoshimatsu, Shoi Shi, Ryo Ueno, Motoi Suzuki, and Daisuke Yoneoka. 2022. “Impact of Airline Travel Network on the Global Importation Risk of Monkeypox, 2022.” bioRxiv. https://doi.org/10.1101/2022.09.17.22280060.

Kuo, Pei-Fen, and Chui-Sheng Chiu. 2021. “Airline Transportation and Arrival Time of International Disease Spread: A Case Study of Covid-19.” PloS One 16 (8): e0256398.

Lai, Shengjie, Isaac I. Bogoch, Nick W. Ruktanonchai, Alexander Watts, Xin Lu, Weizhong Yang, Hongjie Yu, Kamran Khan, and Andrew J. Tatem. 2022. “Assessing Spread Risk of COVID-19 within and beyond China in Early 2020.” Data Science and Management 5 (4): 212–18.

Liao, Qiuyan, Benjamin Cowling, Wing Tak Lam, Man Wai Ng, and Richard Fielding. 2010. “Situational Awareness and Health Protective Responses to Pandemic Influenza A (H1N1) in Hong Kong: A Cross-Sectional Study.” PloS One 5 (10): e13350.

Mathieu, Edouard, Fiona Spooner, Saloni Dattani, Hannah Ritchie, and Max Roser. 2022. “Monkeypox.” Our World in Data, May. https://ourworldindata.org/monkeypox?utm_source=delta%20optimist&utm_campaign=delta%20optimist%3A%20outbound&utm_medium=referral.

Meides, Marco. n.d. “The OpenSky Network - Free ADS-B and Mode S Data for Research.” Accessed September 13, 2022. https://opensky-network.org/.

Menkir, Tigist F., Taylor Chin, James A. Hay, Erik D. Surface, Pablo M. De Salazar, Caroline O. Buckee, Alexander Watts, et al. 2021. “Estimating Internationally Imported Cases during the Early COVID-19 Pandemic.” Nature Communications 12 (1): 311.

“Monkeypox 2022 Global Epidemiology; Report 2022-09-23.” n.d. Accessed January 31, 2023. https://www.monkeypox.global.health/.

“Monkeypox Outbreak.” n.d. Accessed November 7, 2022. https://www.who.int/emergencies/situations/monkeypox-oubreak-2022.

“Monkeypox - United Kingdom of Great Britain and Northern Ireland.” n.d. Accessed November 7, 2022. https://www.who.int/emergencies/disease-outbreak-news/item/2022-DON381.

“Multi-Country Monkeypox Outbreak in Non-Endemic Countries.” n.d. Accessed November 27, 2022. https://www.who.int/emergencies/disease-outbreak-news/item/2022-DON385.

Nakamura, Hiroki, and Shunsuke Managi. 2020. “Airport Risk of Importation and Exportation of the COVID-9 Pandemic.” Transport Policy 96 (September): 40–47.

National Immunization Advisory Committee (NIAC) Technical Working Group (TWG), Influenza Vaccination TWG. 2021. “[Technical guidelines for seasonal influenza vaccination in China (2021-2022)].” Zhonghua liu xing bing xue za zhi = Zhonghua liuxingbingxue zazhi 42 (10): 1722–49.

Nature. 2022. “Monkeypox: Wealthy Countries Must Avoid Their COVID-19 Mistakes.” “Open Data @ OurAirports.” n.d. Accessed September 13, 2022. https://ourairports.com/data/.

Pine, Ray, and Bob McKercher. 2004. “The Impact of SARS on Hong Kong’s Tourism Industry.” International Journal of Contemporary Hospitality Management 16 (2): 139–43.

Reuters. 2022a. “Hong Kong Discovers First Case of Monkeypox,” September 6, 2022. https://www.reuters.com/world/asia-pacific/hong-kong-discovers-first-case-monkeypox-2022-09-06/.

Reuters. 2022b. “Hong Kong to Start Monkeypox Vaccination on October 5,” September 21, 2022. https://www.reuters.com/world/asia-pacific/hong-kong-start-monkeypox-vaccination-october-5-2022-09-21/.

Russell, Timothy W., Joseph T. Wu, Sam Clifford, W. John Edmunds, Adam J. Kucharski, Mark Jit, and Centre for the Mathematical Modelling of Infectious Diseases COVID-19 working group. 2021. “Effect of Internationally Imported Cases on Internal Spread of COVID-19: A Mathematical Modelling Study.” The Lancet. Public Health 6 (1): e12–20.

Shi, Shoi, Shiori Tanaka, Ryo Ueno, Stuart Gilmour, Yuta Tanoue, Takayuki Kawashima, Shuhei Nomura, Akifumi Eguchi, Hiroaki Miyata, and Daisuke Yoneoka. 2020. “Travel Restrictions and SARS-CoV-2 Transmission: An Effective Distance Approach to Estimate Impact.” Bulletin of the World Health Organization 98 (8): 518–29.

Silverstein, W. K., L. Stroud, G. E. Cleghorn, and J. A. Leis. 2020. “Wu JT, Leung K, Leung GM. Nowcasting and Forecasting the Potential Domestic and International Spread of the 2019-nCoV Outbreak Originating in Wuhan, China: A Modelling Study. Lancet 2020; 395: 689--97—In This Article.” The Lancet 395: 689–97.

Talbot, François. 2022. “The Impact of COVID-19 on Airports—and the Path to Recovery.” ACI World (blog). October 6, 2022. https://aci.aero/2022/10/06/the-impact-of-covid-19-on-airports-and-the-path-to-recovery/.

Thornhill, John P., Sapha Barkati, Sharon Walmsley, Juergen Rockstroh, Andrea Antinori, Luke B. Harrison, Romain Palich, et al. 2022. “Monkeypox Virus Infection in Humans across 16 Countries - April-June 2022.” The New England Journal of Medicine 387 (8): 679–91.

“Tourism Statistics.” n.d. Discover Hong Kong. Accessed October 31, 2022. https://www.discoverhongkong.com/eng/hktb/newsroom/tourism-statistics.html.

Viboud, Cécile, Lone Simonsen, and Gerardo Chowell. 2016. “A Generalized-Growth Model to Characterize the Early Ascending Phase of Infectious Disease Outbreaks.” Epidemics 15 (June): 27–37.

Wang, Lin, and Joseph T. Wu. 2018. “Characterizing the Dynamics Underlying Global Spread of Epidemics.” Nature Communications 9 (1): 218.

Wang, Shuqi, Fengdi Zhang, Zhilu Yuan, Mingda Xu, Zhen Wang, Chao Gao, Renzhong Guo, and Zhanwei Du. 2022. “Serial Intervals and Incubation Periods of the Monkeypox Virus Clades.” Journal of Travel Medicine 29 (8): taac105.

“WHO Director-General Declares the Ongoing Monkeypox Outbreak a Public Health Emergency of International Concern.” n.d. Accessed November 7, 2022. https://www.who.int/europe/news/item/23-07-2022-who-director-general-declares-the-ongoing-monkeypox-outbreak-a-public-health-event-of-international-concern.

“World Population Prospects - Population Division - United Nations.” n.d. Accessed September 15, 2022. https://population.un.org/wpp/Download/Standard/CSV/.

Xu, Mingda, Zhanwei Du, Songwei Shan, Xiaoke Xu, Yuan Bai, Peng Wu, Eric H. Y. Lau, and Benjamin J. Cowling. 2022. “RiskEstim: A Software Package to Quantify COVID-19 Importation Risk.” Frontiers of Physics 10. https://doi.org/10.3389/fphy.2022.835992.

Zhang, Lei, Lu Zhang, Li Lai, Zhanwei Du, Yuling Huang, Jianming Su, Canglang Wu, Shujuan Yang, and Peng Jia. 2022. “Risk Assessment of Imported COVID-19 in China: A Modelling Study in Sichuan Province.” Transboundary and Emerging Diseases, September. https://doi.org/10.1111/tbed.14700.

